# The COMT Val^158^Met Polymorphism is Significantly Associated with Early Onset Preeclampsia in Both African American and Caucasian Mothers

**DOI:** 10.1101/2024.05.01.24306705

**Authors:** Melissa R. Kaufman, Amy E. Hwang, Anthony M. Pickrel, Cassandra M. Gray, Kriti M. Goel, David N. Dhanraj, Jerome L. Yaklic, Rose A. Maxwell, Thomas L. Brown

## Abstract

The aim of this study was to evaluate maternal and infant Val^158^Met polymorphisms of Catechol-O-Methyltransferase (COMT), a reported indicator of preeclamptic risk, in a United States population. Healthy control, early-onset preeclamptic, and late-onset preeclamptic patients were enrolled in this study. Genomic DNA was isolated from mothers and infants via buccal swabs and DNA was genotyped via tetra-primer amplification PCR. Our findings indicate that the COMT genotype was not significantly associated with late-onset PE. While there were no significant differences between African American and Caucasian races, the maternal COMT^Met158Met^ genotype was significantly associated with early-onset preeclampsia in both African Americans and Caucasians when compared to COMT^Val158Val^ or COMT^Val158Met^. These results suggest that the maternal COMT^Met158Met^ genotype may be a risk factor for early-onset PE.

## Introduction

Preeclampsia (PE) is a pregnancy-specific condition and a leading cause of maternal and fetal morbidity and mortality [1-5]. It is characterized by abrupt-onset hypertension accompanied by proteinuria or maternal hepatic, pulmonary, renal, visual, or neurological involvement [6-8]. PE can be stratified into two subtypes, early and late onset. Early-onset PE is often more severe, occurs before 34 weeks of gestation, and usually results in placental abnormalities and fetal growth restriction. Late-onset PE, which occurs after 34 weeks, typically does not result in placental abnormalities or fetal growth restriction [3-5]. In addition, ethnic differences in PE have been reported, with African American women in the United States having a 60% higher rate of developing the condition than Caucasian women [9-10]. Although it is unclear why such a stark racial disparity occurs in these individuals, differences in genetic makeup have been proposed as a possibility [11].

Previous studies have suggested that genetic polymorphisms may identify risk factors for PE [11,12]. One such candidate is catechol-O-methyltransferase (COMT). COMT is an enzyme involved in the degradation of neurotransmitters and catecholamines and is involved in regulating blood flow in tissues such as the brain, kidney, heart and placenta [13]. A predominant single nucleotide polymorphism (SNP rs4680) in the COMT gene is present at codon 158. This polymorphism, COMT^Val158Met^, results in a transition from a G to an A nucleotide and is responsible for changing the amino acid from a Valine (Val) to a Methionine (Met). Three genotypes of COMT^158^ have been reported: Val/Val (GG), Val/Met (GA), and Met/Met (AA). These genotypes have been shown to display high, intermediate, and low levels of enzymatic activity, respectively [14].

Several studies from numerous countries have reported that polymorphic maternal COMT^Val158Met^ is a risk factor for the development of PE in Asian, Southeastern European, South American, and Middle Eastern countries [15-19]. In addition, Roten et al., found that low COMT activity was associated with recurrent PE in Norwegian mothers [20].

In contrast, Pertegel et al., found that fetal COMT^Val158Met^ but not maternal was associated with PE in a Spanish population [21]. Conflicting reports by Zhao et al., did not find any significance differences in Chinese mothers with PE [22] whereas studies by Krnjeta et al., found the COMT low activity genotype (Met/Met) was associated with a decreased risk of early-onset PE in a Serbian population [23]. The reason for these discrepancies is unclear.

These previous studies indicate that maternal COMT^Met158Met^ is associated with PE, particularly the early-onset subtype. However, no studies have reported an analysis of maternal or fetal COMT^158^ polymorphisms in North American or racially disparate populations.

## Materials and Methods

The population under study consisted of African American and Caucasian mothers aged 18-40 and their babies: 29 healthy controls; 19 early-onset preeclampsia (<34 weeks gestation), and 43 late-onset preeclampsia (>34 weeks gestation) (Table 1). All subjects were informed and gave written consent prior to their participation in this study at Miami Valley Hospital, a high-risk maternity referral center in Dayton, Ohio USA. This study was reviewed and approved by the Wright State University Institutional Review Board. Genomic DNA was collected from mothers and babies using buccal swabs. Both sides of the oral cavity were swabbed and then stored at - 80°C until processing (Isohelix). Genomic DNA was isolated from buccal swabs per the manufacturer’s instructions and stored at -20C for PCR analysis.

**Table 1:** Demographic and Clinical Characteristics by Diagnosis.

|  | Control<br>(n=29) | EOPE<br>(n=19) | LOPE<br>(n=43) | p-value |
| --- | --- | --- | --- | --- |
| **Maternal Age\*** | 25.0 ± 4.5 | 27.6 ± 7.1 | 28.2 ± 5.7 | 0.058 |
| **Maternal BMI at Delivery\*** | 32.7 ± 5.5 | 37.0 ± 10.0 | 34.7 ± 10.3 | 0.259 |
| **Maternal Race** |  |  |  | 0.140 |
| African American | 16 (55.2%) | 5 (26.3%) | 18 (41.9%) |  |
| Caucasian | 13 (44.8%) | 14 (73.7%) | 25 (58.1%) |  |
| **Gravida\*** | 2.5 ± 1.7 | 2.9 ± 2.5 | 2.9 ± 2.1 | 0.734 |
| **Parity\*** | 1.4 ± 1.3 | 1.9 ± 2.0 | 1.8 ± 1.8 | 0.538 |
| **History of Preterm Birth** | 0 | 4 (21.1%) | 8 (19.0%) | 0.036 |
| **History of gestational Hypertension** | 0 | 2 (10.5%) | 4 (9.3%) | 0.219 |
| **History of Preeclampsia** | 0 | 4 (21.1%) | 8 (18.6%) | 0.038 |
| **Chronic hypertension** | 2 (6.9%) | 12 (63.2%) | 12 (27.9%) | <0.001 |
| **Chronic diabetes** | 0 | 1 (5.3%) | 5 (11.6%) | 0.144 |
| **Gestational diabetes** | 0 | 1 (5.3%) | 4 (9.3%) | 0.236 |
| **Mode of delivery** |  |  |  | 0.613 |
| Vaginal delivery | 15 (53.6%) | 9 (47.4%) | 26 (60.5%) |  |
| Cesarean delivery | 13 (46.4%) | 10 (52.6%) | 17 (39.5%) |  |
| **Gestational age at delivery (weeks)\*** | 39.4 ± 1.1 | 31.9 ± 1.7 | 37.5 ± 1.7 | <0.001 |
| **Birth weight (g)\*** | 3293.4 ±<br>546.7 | 1592.6 ±<br>543.1 | 2830.0 ±<br>826.7 | <0.001 |
| **Infant Sex** |  |  |  | 0.200 |
| Male | 17 (58.6%) | 12 (63.2%) | 18 (41.9%) |  |
| Female | 12 (41.4%) | 7 (36.8%) | 25 (58.1%) |  |
| **NICU admission** | 1 (3.4%) | 19 (100%) | 15 (34.9%) | <0.001 |
| **NICU length of stay** | --* | 26.6 ±14.3 | 15.1 ± 18.6 | 0.053 |
| **Apgar 1 minute\*** | 7.9 ± 0.8 | 7.3 ± 1.3 | 7.0 ± 1.9 | 0.069 |
| **Apgar 5 minute\*** | 8.9 ± 0.3 | 8.6 ± 0.6 | 8.6 ± 1.1 | 0.215 |

\*only 1 case had a NICU stay; t-test results presented. \* Mean +/- Standard Deviation.

Analysis of COMT^Val158Met^ was performed via genomic PCR and was carried out using a tetra-primer amplification refractory mutation system PCR analysis with modifications [Figure 1; 19]. The primers used were as follows: P1 5’-CCAACCCTGCACAGGCAAGAT-3’ (forward), P2 5’-CAAGGGTGACCTGGAACAGCG-3’ (Reverse), P3 5’-CGGATGGTGGATTTCGCTGACG-3’ (forward), P4 5’-TCAGGCATGCACACCTTGTCCTTTAT-3’ (Reverse) [24]. PCR reactions contained nuclease free water, 20-40ng of isolated genomic DNA, COMT P1 - P4 Primer Mix (1:1:1:1 ratio), and Quick Load Taq 2x Master Mix in a final volume of 40ul. PCR was performed under the following conditions: one cycle at 94°C for 4 minutes, 27 cycles of [denaturation at 94°C for 30 seconds, amplification at 65°C for 30 seconds, and elongation at 72°C for 20 seconds], with a final extension cycle at 72°C for 5 minutes. The PCR products were electrophoresed on a 1.5% agarose gel and the DNA was stained using ethidium bromide. Characteristic COMT genotype PCR products were identified as Val/Val (626bp and 451bp), Val/Met (626bp, 451bp, 222bp) and Met/Met (626bp, 222bp) [Figure 1, 24].

**Figure 1:**
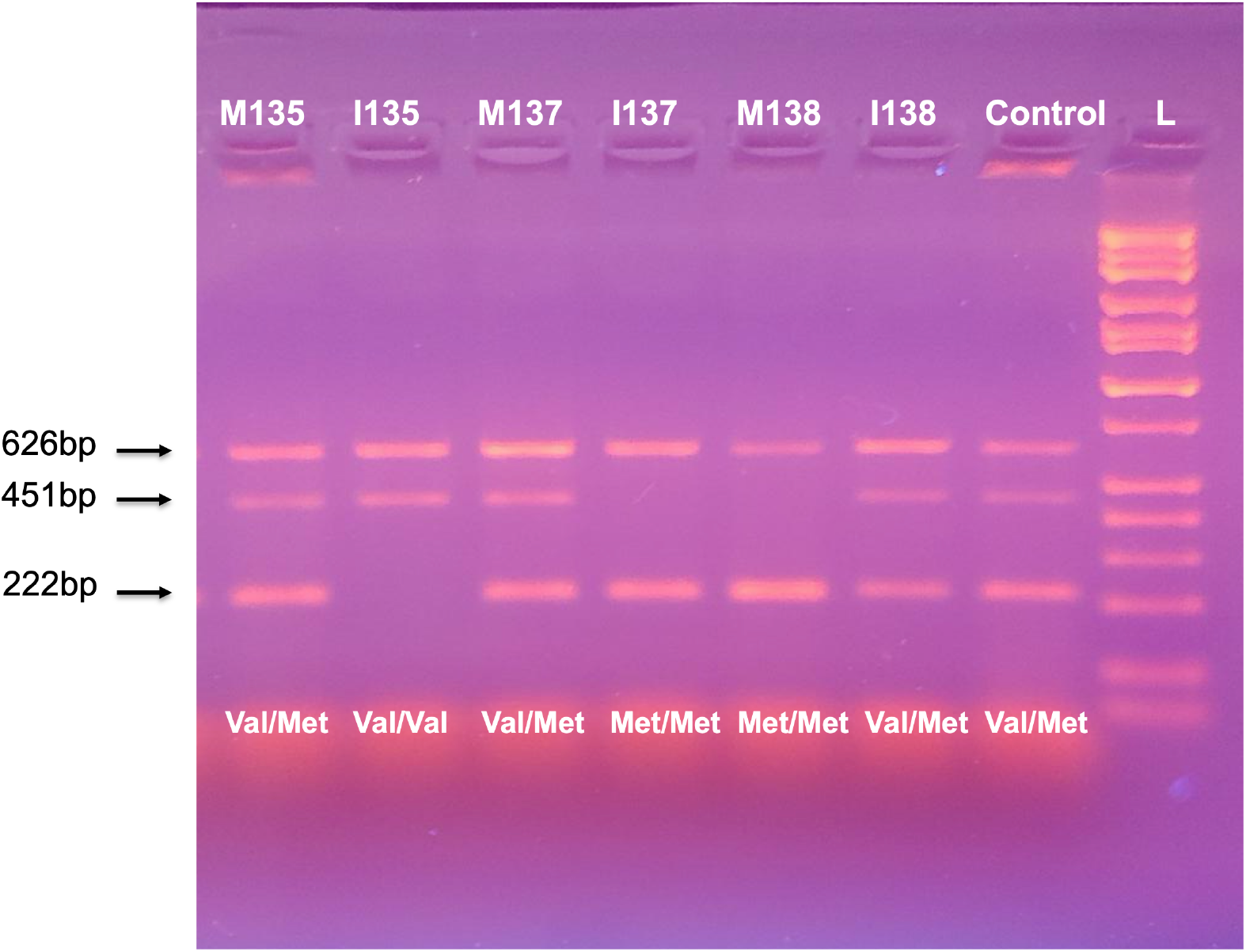
Tetra-Primer Amplification Genomic PCR analysis of COMT158 polymorphisms. Representative figure of COMT158 polymorphisms. All genomic samples have a control 626 base pair (bp) band. The 222bp band indicates the COMTMet158Met genotype (Homozygous AA). The 451 bp band indicates the COMTVal158Val genotype (Homozygous GG). The presence of the 222bp and 451bp indicates the COMTVal158Met genotype (Heterozygous GA). **M** indicates maternal samples. **I** indicates infant samples. The number following the letter is the subject number. Control is a heterozygous positive sample. **L** is the DNA molecular weight ladder.

Statistics were analyzed with separate logistic regression models for all comparisons using SPSS version 29.0 (IBM; Armonk, NY). Odds ratios (OR) and 95% confidence intervals (CI) were used to evaluate variables in the prediction models which included covariates of interest (delivery BMI, maternal race, infant sex, and chronic hypertension). A p< 0.05 was used to determine statistical significance.

## Results and Discussion

The demographic and clinical characteristics of the PE groups and healthy pregnancies are described in Table 1. The results indicated that COMT genotypes in PE pregnancies were not significantly different between African American and Caucasian mothers when comparing parental race (Table 2 and 3). In addition, no significant differences among PE pregnancies were found between COMT genotypes when comparing infant sex (Table 2 and 3). Furthermore, COMT genotypes were not associated with the development of late onset PE (Table 2).

**Table 2:** Regression Model Predicting Late-onset PE vs Healthy Control Grouping.

| Variable | OR (95% CI) | p-value |
| --- | --- | --- |
| Maternal Genotype | 0.54 (0.25-1.20) | 0.13 |
| Delivery BMI | 1.04 (0.96-1.12) | 0.34 |
| Parental Race | 0.52 (0.18-1.53) | 0.23 |
| Sex of Infant | 0.49 (0.17-1.42) | 0.19 |
| Chronic Hypertension | 7.36 (1.31-41.29) | 0.02* |

**Table 3:** Regression Model Predicting Early-onset PE vs Healthy Control Grouping.

| Variable | OR (95% CI) | p-value |
| --- | --- | --- |
| Maternal Genotype | 0.06 (0.01-0.43) | 0.005* |
| Delivery BMI | 1.11 (0.96-1.29) | 0.170 |
| Parental Race | 0.14 (0.02-1.38) | 0.090 |
| Sex of the Infant | 7.49 (0.55-100.90) | 0.130 |
| Chronic hypertension | 184.60 (6.25-5449.93) | 0.003* |

Our studies did not find a significant association in early-onset or late-onset PE with body mass index (BMI) at delivery but did find significance with preexisting chronic hypertension (Table 2 and 3). In addition, the maternal genotype in both African Americans and Caucasians was significantly associated with early-onset PE (Table 3). Further analysis indicated that the COMT^Met158Met^ genotype, known to be associated with low enzyme activity, was significantly associated in early-onset PE when compared to COMT^Val158Val^, which expresses high enzyme levels, as well as the combined COMT^Val158Val^/COMT^Val158Met^ genotypes (Table 4, Models 1 and 2). No significant differences were noted between COMT^Val158Val^ and COMT^Val158Met^ genotypes (Table 4, Model 4) or COMT^Val158Met^ and ^Met158Met^ (Table 4, Model 3). Our results indicate that the maternal COMT^Met158Met^ genotype is significantly associated with the development of early-onset PE compared to the COMT^Val158Met^ and COMT^Val158Val^ genotypes.

**Table 4:**
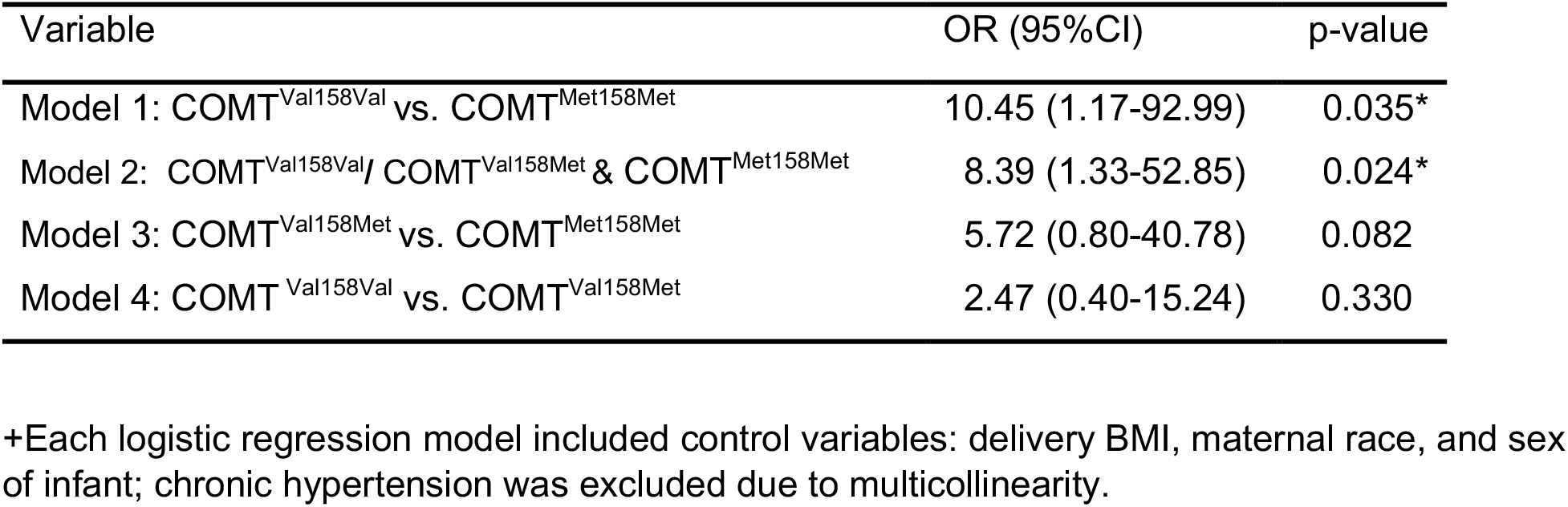
Odds Ratios for Maternal Genotype in Logistic Regressions Predicting Early-onset PE vs Healthy Control Grouping+.

Our study provides several findings not previously reported. We used noninvasive buccal swab analysis to determine if the COMT^Val158Met^ genotype was associated with racial disparity, genomic DNA from African American and Caucasian mothers and infants were analyzed. This patient population was from the midwestern United States. We also assessed infant COMT genotypes to determine if an association existed from being born from a PE pregnancy in African Americans and Caucasians. We further evaluated COMT genotypes to determine if they would be associated with early-onset or late-onset PE subtypes. Our results indicate that the maternal COMT^Met158Met^ genotype is significantly associated with the development of early-onset PE compared to the COMT^Val158Met^ and COMT^Val158Val^ genotypes. In addition, the absence of an association between COMT^Val158Met^ polymorphisms and race suggests that other factors may account for ethnic differences and the increased frequency of PE. Our studies suggest further analysis of increased sample sizes is warranted. Overall, our findings suggest that the maternal COMT^Met158Met^ genotype may serve as an easily identifiable genomic risk factor for early-onset PE.

## Data Availability

All data produced in the present work are contained in the manuscript

## Acknowledgements

We would like to thank the mothers and babies at Miami Valley Hospital in Dayton, Ohio for graciously participating in this research and Mike Bottomley and Summer A. Paschal, for statistical advice.

